# Uncertainty Quantification of Central Canal Stenosis Deep Learning Classifier from Lumbar Sagittal T2-Weighted MRI

**DOI:** 10.1101/2025.10.24.25338153

**Authors:** Adriana Brenzikofer, Maria Monzon, Fabio Galbusera, Zina-Mary Manjaly, Andrea Cina, Catherine R. Jutzeler

**Author notes:** **Correspondence** Maria Monzon, Department of Health Sciences and Technology, ETH Zurich, Zurich, 8092, Switzerland. **Funding information** Strategic Focus Area “Personalized Health and Related Technologies (PHRT)”, ETH Domain (Swiss Federal Institute of Technology), Grant Number: 380; Ambizione Grant. Grant number: #PZ00P3_186101, CRJ and the Schulthess Klinik Research Fund (FG). Equally contributing authors (Adriana Brenzikofer, Maria Monzon). Shared last authorship (Andrea Cina, Catherine R. Jutzeler).

## Abstract

**Background:** Accurate assessment of the severity of central canal stenosis (CCS) on lumbar spine MRI is critical for clinical decision-making. We evaluated deep learning models for automated CCS grading on sagittal T2-weighted MRI, focusing on uncertainty quantification to improve clinical reliability.

**Methods:** Using a retrospective cohort from the LumbarDISC dataset (1,974 patients), we compared multiple deep learning architectures for three-level CCS classification (normal / mild, moderate, severe). To assess model confidence, Monte Carlo (MC) dropout and Test Time Augmentation (TTA) techniques were applied to quantify prediction uncertainty.

**Results:** The fine-tuned Spinal Grading Network (SGN) achieved a balanced accuracy of 79.4% and a macro F1 score of 68.8%, with per-class accuracies of 71.3% for moderate and 78.5% for severe stenosis. MC dropout revealed an increase in uncertainty predominantly in moderate and severe cases, while TTA uncertainty was higher for mild stenosis.

**Conclusion:** DL-based CCS grading demonstrates potential to assist radiologists by providing rapid, standardized evaluations. Incorporating uncertainty quantification offers a safeguard to flag ambiguous cases, thus supporting clinical trust and facilitating safer integration of AI tools into the interpretation of spine MRI.

## 1 INTRODUCTION

Low back pain (LBP) remains the leading cause of years lived with disability worldwide, often leading to diminished mobility and independence [1]. One significant aspect of LBP is lumbar spinal stenosis (LSS). This is characterized by narrowing of the spinal canal, which can compress the spinal cord and/or nerve roots, often leading to pain, numbness and disability [2]. The diagnosis of LSS is based on clinical history, physical examination, and Magnetic Resonance Imaging (MRI) of the lumbar spine [3]. However, interpreting MRI scans is complex and time consuming, involving multiple slices and intervertebral levels. This can lead to inconsistent classification among radiologists [4] and delayed diagnoses. In addition, clinical presentation and radiological presentation do not necessarily correlate.

Deep learning (DL) models have emerged as a promising solution to address these gaps, with the potential to assist clinicians by automating the detection and grading of LSS, as well as other pathological findings observed in radiological imaging. Several DL models have been developed to detect and classify LSS [5, 6, 7, 8, 9, 10]. Among these, SpineNetV2 [7, 11] is considered the most advanced model. It was developed to detect vertebral levels and intervertebral discs (IVD) and grade degenerative findings, including central canal stenosis (CCS), a form of lumbar spinal stenosis (LSS) characterized by compression or inflammation of the spinal cord. SpineNetV2 has also been externally validated in different clinical datasets [12, 13, 14, 15]. However, a major barrier to the clinical adoption of DL models is to ensure their trustworthiness; without quantifying prediction uncertainty, these models remain difficult to trust or implement in clinical practice [16].

To address this challenge, it is essential to understand that uncertainty in DL models can be broadly classified into epistemic and aleatoric types [17]. Epistemic uncertainty, often referred to as model uncertainty, arises from limitations in the model’s knowledge that are typically due to insufficient diverse training data or inadequate model architecture [18]. Monte Carlo (MC) dropout [19] is widely used to estimate epistemic uncertainty. Aleatoric uncertainty is the inherent randomness or irreducible noise in the data that cannot be resolved by adding more training samples [17]. This type of uncertainty in LSS MRI can arise from motion artifacts, anatomical variability, and ambiguous tissue boundaries with overlapping signal intensities that obscure the spinal canal. Test-Time Augmentation (TTA) is a common method to quantify this data-inherent uncertainty [20]. Although uncertainty estimation methods such as MC dropout and TTA are used in medical imaging [21, 22], their effectiveness to improve model reliability and help clinical evaluations is underexplored.

In this study, we developed a robust DL model for classifying CCS severity [23] from sagittal MRI T2-weighted scans, emphasizing the critical role of uncertainty quantification in clinical settings. Although calibration analysis assessed the alignment between predicted probabilities and real-world outcomes, calibrated probabilities alone do not fully capture predictive uncertainty [24, 25]. To better quantify uncertainty, we used MC dropout and TTA, which capture predictive variability beyond standard softmax scores. The main objectives were to (1) compare different DL architectures to classify CCS severity, (2) quantify prediction uncertainty using MC dropout and TTA, and (3) investigate the use of majority voting in the MC and TTA outputs to improve model performance. An overview of the full methodology and uncertainty estimation pipeline is provided in Fig. 1. This approach aims to address clinical needs by providing automated CCS classification accompanied by confidence measures, thereby supporting better decision making through explicit awareness of uncertainty.

**FIGURE 1.**
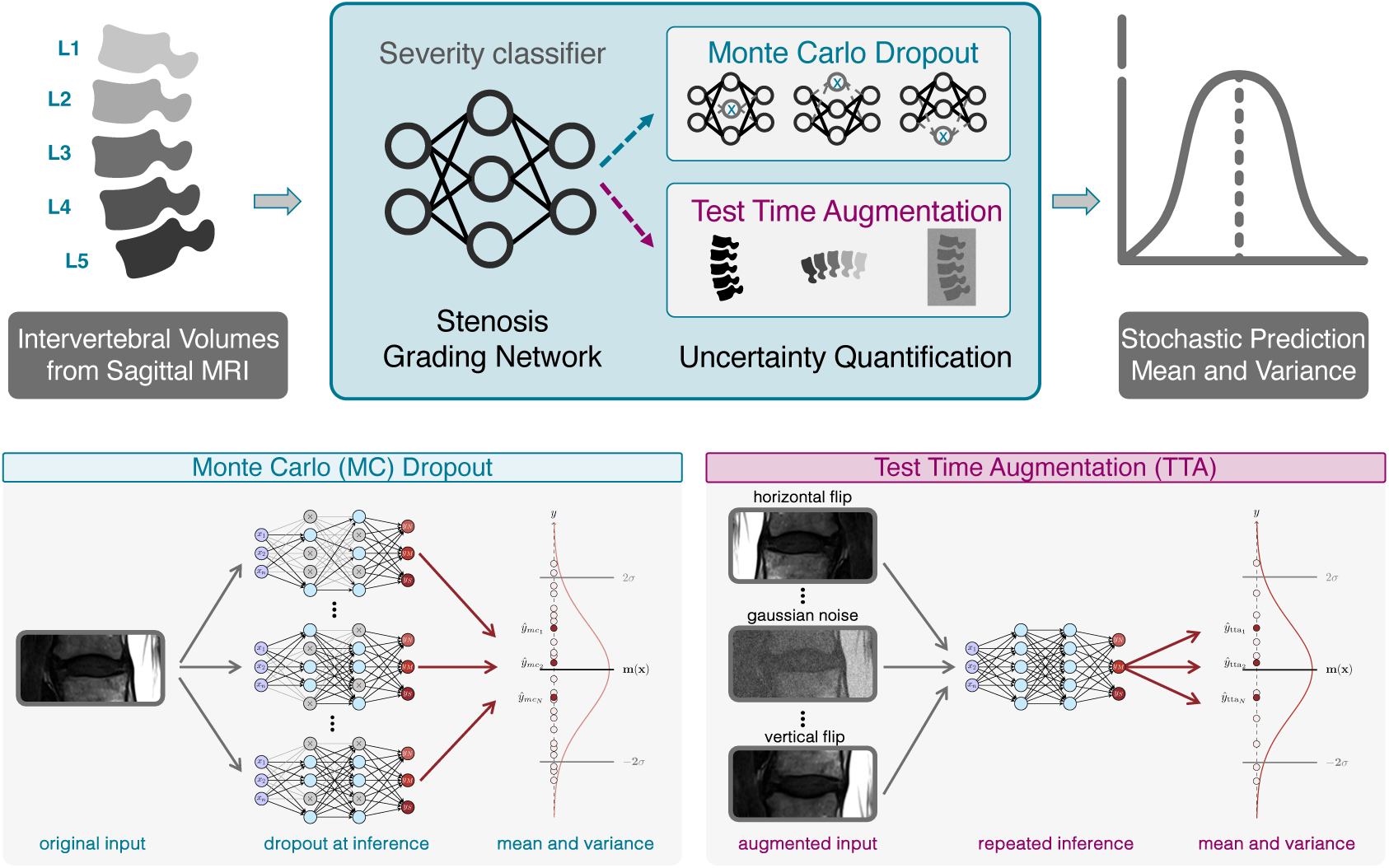
Overview of the uncertainty quantification pipeline for central canal stenosis grading. The pipeline extracts intervertebral volumes from sagittal T2-weighted lumbar spine MRIs, from intervertebral levels L1-L2 to L5-S1. A fine-tuned Spine Grading Network (SGN), adapted from the pre-trained SpineNetV2 ResNet-34 architecture incorporating dropout predicts severity of the stenosis (normal/mild, moderate, severe). To quantify uncertainty,two methods are employed to generate stochastic predictions: Test-Time Augmentation (TTA) to estimate data-inherent aleatoric uncertainty and Monte Carlo (MC) Dropout to estimate model-based epistemic uncertainty. Multiple forward passes through the network yield a distribution of predictions for each input volume.

## 2 MATERIALS AND METHODS

### 2.1 Dataset preprocessing

The data set used in this study consisted of sagittal T2-weighted MRI scans of the lumbar spine from the LumbarDISC dataset [26], comprising 1,974 patients across eight international medical centers. Radiologists evaluated and annotated five disc levels (L1-L2 to L5-S1) per patient using a three-tier classification system: normal/mild, moderate, and severe stenosis. Following SpineNetV2 pipeline [7], pre-processing included slice-wise normalization to the range [0, 1], with the median intensity of adjacent vertebral bodies that comprise the IVD level set to 0.5. Intervertebral volumes were extracted using the SpineNetV2 detection module, which automatically localises each disc level and centres the 9-slice sagittal stack on the mid-sagittal plane, consistently capturing the central spinal canal across scanners and imaging protocols. The preprocessed volumes of the final input had dimensions of 9 × 112 × 224 voxels (num slices × height × width).

### 2.2 CCS classification model

SpineNetV2 was originally trained on the Genodisc dataset [11]. It combines vertebral detection with different classification subnetworks to predict specific degenerative conditions, including the four severity grades of CCS: normal, mild, moderate, and severe based on the Lee classification system [23]. We adapted the SpineNetV2 stenosis classification subnetwork to predict CCS severity according to the three-tier RSNA challenge system: normal / mild, moderate and severe stenosis. Specifically, the final layer of the ResNet34-based SpineNetV2 Grading Network (SGN) was adapted to predict three classes instead of four. Adapting a pre-trained network to a new, related task and retraining only part of the network is known as fine-tuning. In addition, we applied dropout regularization [27] to various layers of the adapted SGN. Dropout regularization is a technique used during neural network training in which certain neurons are randomly deactivated to encourage the model to generalise better and avoid over-reliance on specific features. In our model, dropout was applied between layer 4 and the average pooling layer, as well as before the final layer. This step was necessary to enable MC dropout, which allows the model to estimate predictive uncertainty. To benchmark the performance of the fine-tuned SGN, we also evaluated three alternative DL architectures: DenseNet [28], ResNet [29], and EfficientNet [30], all implemented within the Medical Open Network for Artificial Intelligence (MONAI) framework [31].

For the development of DL models, we divided our MRI data into 70% for model training, 15% for validation during development, and 15% for final testing, ensuring that a single patient did not appear in multiple splits, avoiding data leakage and ensuring that the proportions of the three stenosis severity grades were similar in all sets. We evaluated the model on the test set using balanced accuracy to address class imbalance, and macro F1 score to capture the trade-off between precision and recall across all stenosis severity grades. Additionally, we computed the confusion matrix and, for each class, calculated precision (the proportion of cases predicted as severe that are truly severe) and per-class accuracy (the proportion of true severe cases correctly identified). To assess regional performance differences, we also conducted class-specific evaluations at each level of the intervertebral disc (IVD).

### 2.3 Uncertainty estimation

We performed an analysis of the calibration of the model’s output probabilities using the Expected Calibration Error (ECE) per class [32]. An ECE of zero means that the probabilities are perfectly calibrated and can be used as a measure of uncertainty. However, since the output probabilities of the DL model are often miscalibrated [24], to enhance the clinical applicability of our top performing CCS classification model, we implemented two uncertainty estimation methods: MC dropout [19] and TTA [20].

MC dropout offers a quantitative evaluation of the epistemic uncertainty of the DL model by maintaining random neuron deactivation during prediction [27]. This technique extends conventional dropout regularization beyond training by maintaining stochastic neuron deactivation during inference, generating multiple predictions from identical input data. The variance of the resulting predictions directly correlates with the uncertainty of the model: higher variance signifies greater uncertainty of the model and, consequently, reduced confidence in any individual prediction, highlighting cases requiring radiologist review. We implemented 25 dropout iterations, as it was observed that, beyond 20 iterations, the predictive uncertainty exhibited minimal variation [33].

To quantify aleatoric uncertainty, which arises from inherent data variability, we implemented TTA [20]. In TTA, augmentations serve as controlled input perturbations rather than realistic clinical simulations. Therefore, anatomical realism is not a prerequisite for valid aleatoric uncertainty estimation. In each TTA iteration, we applied transformations such as horizontal flipping, rotation, scaling, or the addition of Gaussian noise to the image (see appendix Fig. 7). Running the model on these modified scans generates a prediction distribution that quantifies uncertainty through the predictive mean and variance. The variance of the multiple runs measures how much these predictions vary from one another, serving as a direct quantification of aleatoric uncertainty.

For both MC dropout and TTA, we computed the mean and standard deviation of predicted probabilities associated with the ground-truth class across all iterations.Higher variance indicates greater uncertainty. As a complementary uncertainty measure, we also computed predictive entropy from the mean class probability distribution across repeated stochastic predictions. We conducted a comparative visualization of uncertainty estimations between MC and TTA methods. The predictions were categorized according to the severity of the stenosis (normal / mild, moderate, severe) and further filtered by true labels to assess the alignment between the predicted results and actual diagnoses for each method.

This approach ensures consistency and interpretability, as it allows one to directly quantify uncertainty in the model’s prediction with respect to the correct diagnosis. The low and high uncertainty cases were identified by analyzing the distribution of standard deviations of predicted probabilities for the true class, computed separately for each method and each class. Images with standard deviations above the 75*^th^* percentile were marked as highly uncertain, while those below the 25*^th^* percentile were considered low uncertainty. Confidence, in contrast, was evaluated based on the mean predicted probability for the true class: higher values indicate more confident predictions. It is important to distinguish between confidence and uncertainty. Confidence reflects the model’s belief in a particular prediction, while uncertainty captures how stable and consistent that belief is across multiple stochastic iterations.

#### 2.3.1 Uncertainty-Driven Consensus for Reliable CCS Grading

In addition to using TTA and MC for uncertainty quantification, we combined both methods with majority voting, as TTA with majority voting has shown significant performance improvements in medical image analysis [34]. Building on this, TTA and MC dropout were further leveraged by aggregating the predictions through majority voting. For each method, we systematically combined the predictions in iterations (25 for MC dropout, 9 for TTA) to derive more reliable stenosis classifications. The majority voting procedure summed class-specific predictions across iterations, with the class receiving the highest vote count designated as the final prediction.

## 3 RESULTS

### 3.1 Data Analysis

This study retrospectively used T2-weighted sagittal magnetic resonance imaging of the lumbar spine of patients 1,974 from the LumbarDISC data set [26]. The resolution of the scans ranges from 0.27 x 0.27 x 3 mm to 1.18 x 1.18 x 6 mm, the mean number of slices 17.01 ± 2.8.

After applying the SpineNetV2 detection module, 9,830 volumes of intervertebral disc (IVD) were extracted for the training of the CCS network. The anatomical distribution revealed severe stenosis mainly in L4-L5 and moderate in L3-L4. Figure 2 shows sample sagittal slices of IVD volumes at level L3-L4 for the three classes of severity of the stenosis. The data set exhibited a substantial class imbalance, with 88% (n=8,630) of IVD volumes classified as normal/mild stenosis, while moderate and severe stenosis represented only 7% (n=732) and 5% (n=468) of the dataset, respectively.

**FIGURE 2.**
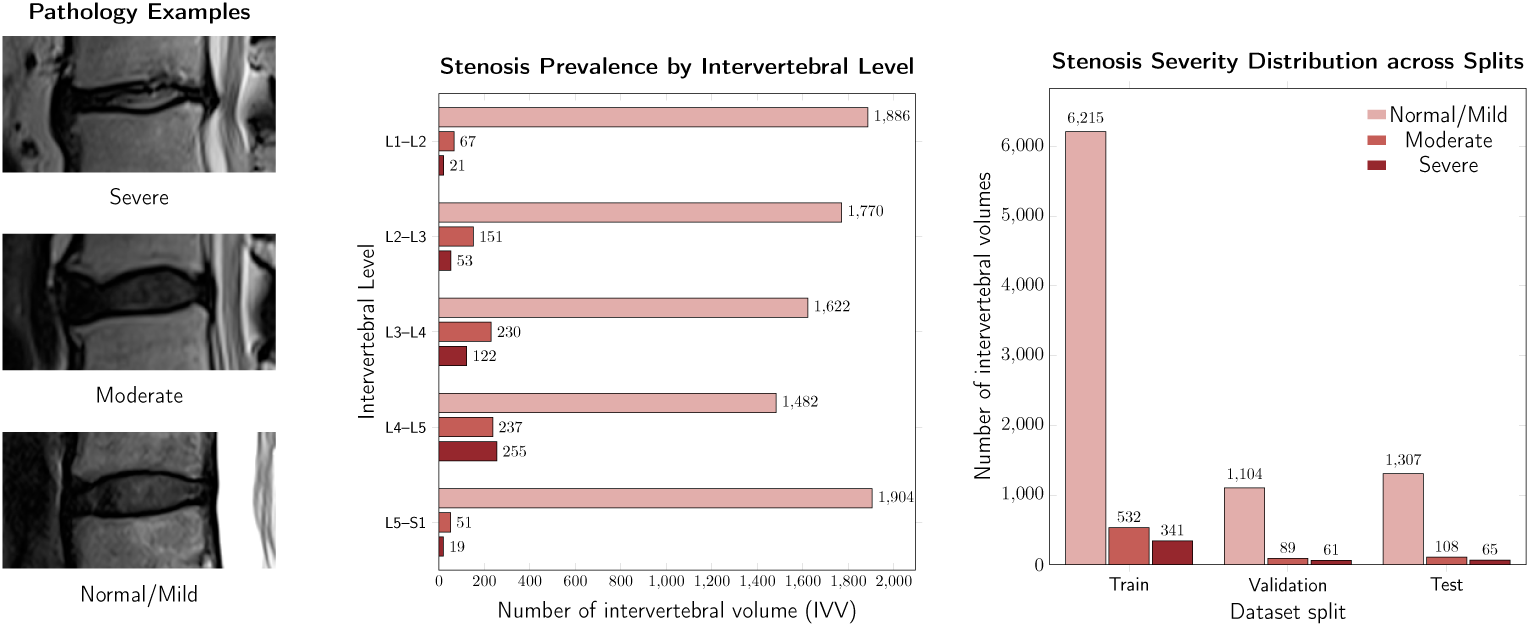
*(a)* Examples of Central Canal Stenosis Severity. *(b)* Distribution of stenosis severity for each of intervertebral level. *(c)* Distribution of stenosis severity of intervertebral disc (IVD) labels across train, validation, and test splits and individual patients per split.

### 3.2 CCS classification model performance

In the comprehensive evaluation of the five deep learning architectures for lumbar spinal stenosis classification, the fine-tuned SGN demonstrated superior performance, in terms of macro F1 (68.8%) and balanced accuracy (79.4%). Consequently, this model was selected as the best performing architecture for further analysis and uncertainty estimation procedures. The remaining deep learning architectures exhibited comparatively lower performance in key evaluation metrics. DenseNet achieved the second highest performance with a balanced accuracy of 69.0% and a macro F1 score of 68.7%, closely followed by ResNet (balanced accuracy: 65.4%, macro F1: 65.6%) and Efficient-Net (balanced accuracy: 65.6%, macro F1: 66.3%). In particular, the original SpineNetV2 architecture demonstrated the poorest overall performance, with the lowest macro F1 score (59.6%) and balanced accuracy (55.4%) among all evaluated models.

Class-specific analysis (Tab. 1) showed different performance patterns in severity categories of stenosis. Detailed confusion matrices for all five architectures are provided in Supplementary Fig. 6. The original SpineNetV2 model had variable performance, with an accuracy of 98.2% for normal / mild stenosis, but only 20.4% and 47.7% for moderate and severe stenosis. The fine-tuned SGN showed the highest accuracy: 88.4% for normal / mild, 71.3% for moderate and 78.5% for severe stenosis.

**TABLE 1.** Performance evaluation comparison for lumbar spinal central canal stenosis (CCS) classification.

| Metric (%) | fine-tuned SGN | SpineNetV2 | DenseNet | ResNet | EfficientNet |
| --- | --- | --- | --- | --- | --- |
| Accuracy |  |  |  |  |  |
| Normal/Mild | 88.4 | 98.2 | 96.3 | 96.1 | 96.4 |
| Moderate | 71.3 | 20.4 | 53.7 | 55.6 | 48.1 |
| Severe | 78.5 | 47.7 | 56.9 | 44.6 | 52.3 |
| Balanced Accuracy | 79.4 | 55.4 | 69.0 | 65.4 | 65.6 |
| F1 Score |  |  |  |  |  |
| Normal/Mild | 93.5 | 95.6 | 96.8 | 96.5 | 96.6 |
| Moderate | 45.8 | 27.0 | 48.7 | 49.0 | 44.8 |
| Severe | 67.1 | 56.4 | 60.7 | 51.3 | 57.6 |
| F1 Macro Averaged | 68.8 | 59.6 | 68.7 | 65.6 | 66.3 |
| Precision |  |  |  |  |  |
| Normal/Mild | 99.2 | 93.0 | 97.4 | 97.0 | 96.7 |
| Moderate | 33.8 | 40.0 | 44.6 | 43.8 | 41.9 |
| Severe | 58.6 | 68.9 | 64.9 | 60.4 | 64.2 |
| Weighted Mean Precision | 92.7 | 88.1 | 92.1 | 91.5 | 91.3 |
| Sensitivity |  |  |  |  |  |
| Normal/Mild | 88.4 | 98.2 | 96.3 | 96.1 | 96.4 |
| Moderate | 71.3 | 20.4 | 53.7 | 55.6 | 48.1 |
| Severe | 78.5 | 47.7 | 56.9 | 44.6 | 52.3 |
| Weighted Mean Sensitivity | 86.8 | 90.3 | 91.5 | 90.9 | 90.9 |

Higher values indicate better performance in percentages.

The fine-tuned SGN achieved 99.2% precision for normal/mild cases, indicating high confidence for non-stenotic conditions. The precision for moderate stenosis was low in all models, with the fine-tuned SGN at 33.8% and DenseNet outperforming at 44.6%, indicating a high rate of false positive predictions. For severe stenosis, the original SpineNetV2 had 68.9% precision, while the fine-tuned SGN achieved 58.6%.

### 3.3 Uncertainty Estimation for LSS classification model

The ECEs for all stenosis classes were around 0.15, confirming the poor calibration of the model’s probabilities. The uncertainty quantification analysis of our spinal classification network (SGN) using MC dropout and TTA revealed distinct patterns in model confidence across severity grades of CCS (Figure 8). MC dropout demonstrated elevated uncertainty in clinically critical categories, with predictive variance values of 0.7 for moderate and severe stenosis, compared to 0.3 for normal/mild cases. In contrast, TTA exhibited its highest uncertainty (4.5 variance) in normal/mild classifications, while showing greater confidence in severe stenosis predictions (2.6 variance) and moderate cases (2.2 variance).

Figure 3 illustrates the distribution of predicted probabilities and associated uncertainty (quantified by the standard deviation of the predicted probability for the true class) in the three classification classes for MC dropout and test time augmentation (TTA). For Normal/Mild cases, both methods produced high mean predicted probabilities of 0.97 and 0.88 for MC and TTA respectively, suggesting confident predictions. However, MC dropout exhibited a markedly lower spread (standard deviation) compared to TTA, indicating more consistent and certain predictions for this class. The median standard deviation was 0.031 for MC and 0.19 for TTA. For moderate cases, the mean predicted probabilities were lower overall (0.62 for MC and 0.57 for TTA), suggesting the model was less confident in assigning this label. Interestingly, the standard deviations were relatively low, especially for the MC dropout with a median of 0.088, suggesting that, although the model was not confident in its classification (lower mean), it was consistent (low uncertainty). For severe cases, both methods achieved higher mean predicted probabilities of 0.78 and 0.75 for MC and TTA respectively, but TTA again demonstrated greater variability. MC dropout maintained lower standard deviations (mean 0.075), implying more stable predictions. Overall, TTA consistently exhibited greater predictive variation (higher standard deviation) across all classes, suggesting that MC dropout provides more stable uncertainty estimates, particularly for Normal/Mild and Severe grades. For MC dropout, the thresholds above which a case was considered highly uncertain were 0.09, 0.06, and 0.09 for Normal/Mild, Moderate, and Severe cases, respectively. For TTA, the corresponding thresholds were 0.16, 0.25, and 0.22. The cases were marked as low uncertainty if the standard deviation of the predicted probabilities for the true class was below the following thresholds: 0.06, 0.01, and 0.05 for MC dropout, and 0.09, 0.11, and 0.12 for TTA, corresponding to the Normal/Mild, Moderate, and Severe categories, respectively. Figure 3 additionally presents predictive entropy as a complementary uncertainty measure; additional details on entropy-based analyses and Expected Calibration Error (ECE) are provided in the Supplementary Material (Sections 4 and 10). Misclassified cases, predominantly in the Normal/Mild and Severe categories, exhibited markedly higher predictive entropy than correctly classified cases (Supplementary Fig. 9). To further illustrate these findings, the representative IVD magnetic resonance volumes corresponding to cases with high and low predictive uncertainty for both MC dropout and TTA are provided in Figure 4.

**FIGURE 3.**
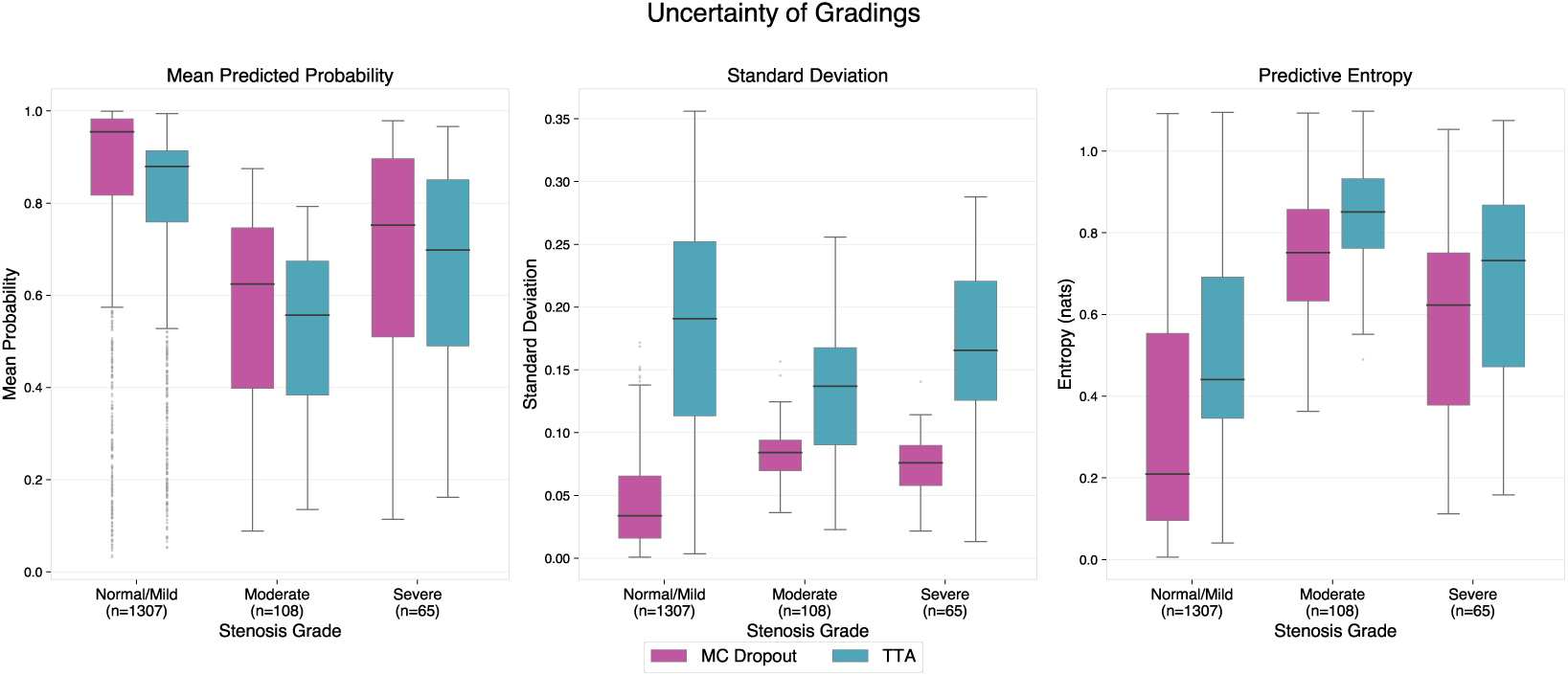
Box plots of uncertainty quantification metrics across Central Canal Stenosis (CCS) severity grades (Normal/Mild, *n* = 1307; Moderate, *n* = 108; Severe, *n* = 65) for MC Dropout (25 stochastic inference passes) and Test-Time Augmentation (TTA, 9 augmented inference passes). **Left**: Mean predicted probability for the assigned severity class, reflecting model confidence. **Centre:** Standard deviation of the predicted class probability across stochastic iterations, quantifying prediction variability. **Right:** Predictive entropy, computed from the mean softmax distribution across stochastic forward passes, providing a complementary measure of overall prediction uncertainty. Across methods, entropy was lowest for Normal/Mild and higher for Moderate and Severe stenosis, consistent with greater diagnostic ambiguity in these classes.

**FIGURE 4.**
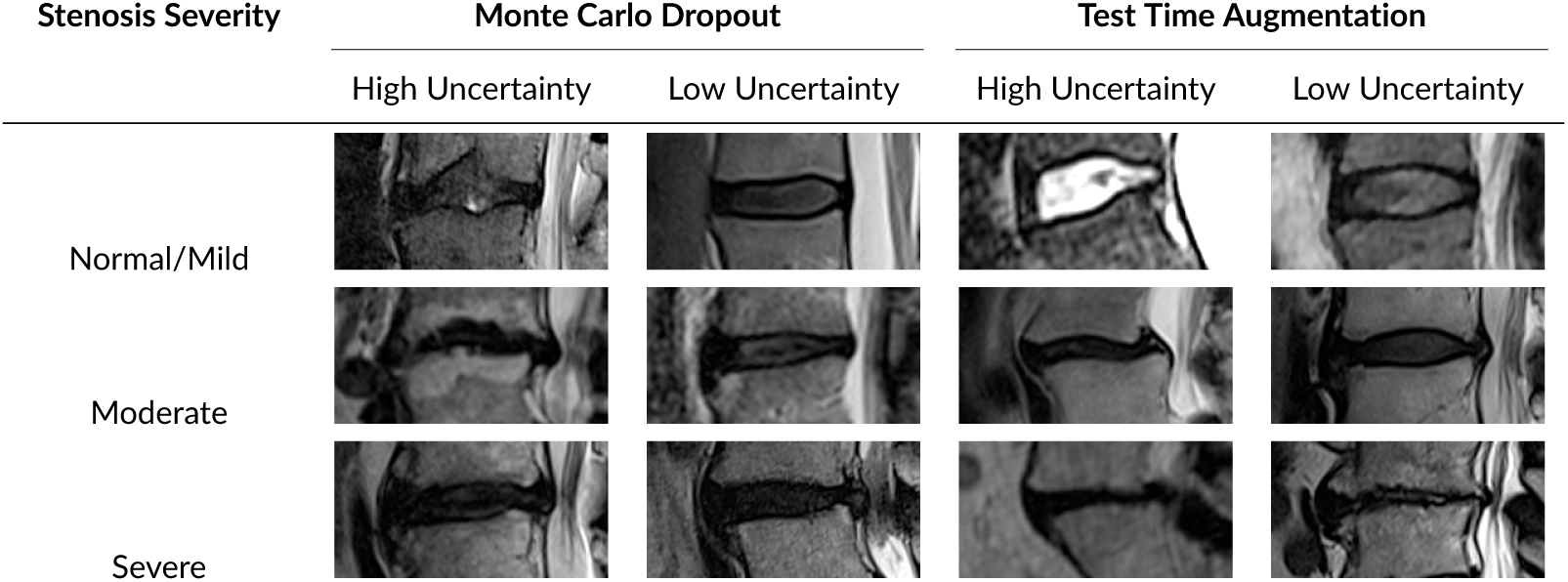
Visualization of high and low uncertainty examples across stenosis severity classes for Monte Carlo (MC) Dropout and Test Time Augmentation (TTA) methods.

These examples highlight that images associated with higher model uncertainty often reflect more degenerated cases or ambiguous anatomical features, emphasizing the practical value of uncertainty quantification for clinical decision support.

#### 3.3.1 Uncertainty-Aware Consensus Performance in Spinal Stenosis Grading

The confusion matrix of uncertainty quantification methods using majority voting is detailed in Fig 5. The application of majority voting to the dropout of MC improved the macro F1 score from 68.8% to 69.8%, indicating enhanced classification consistency, although the balanced accuracy decreased slightly to 77.4% compared to the baseline SGN performance of 79.4%. In contrast, the majority vote applied to TTA did not produce performance improvements, with a macro F1 score of 62.7% and a balanced accuracy remaining at 77.4%. These results suggest that the predictions of the consensus of MC dropout may increase the performance in the classification of CCS, while the consensus of the TTA does not significantly affect overall performance.

**FIGURE 5.**
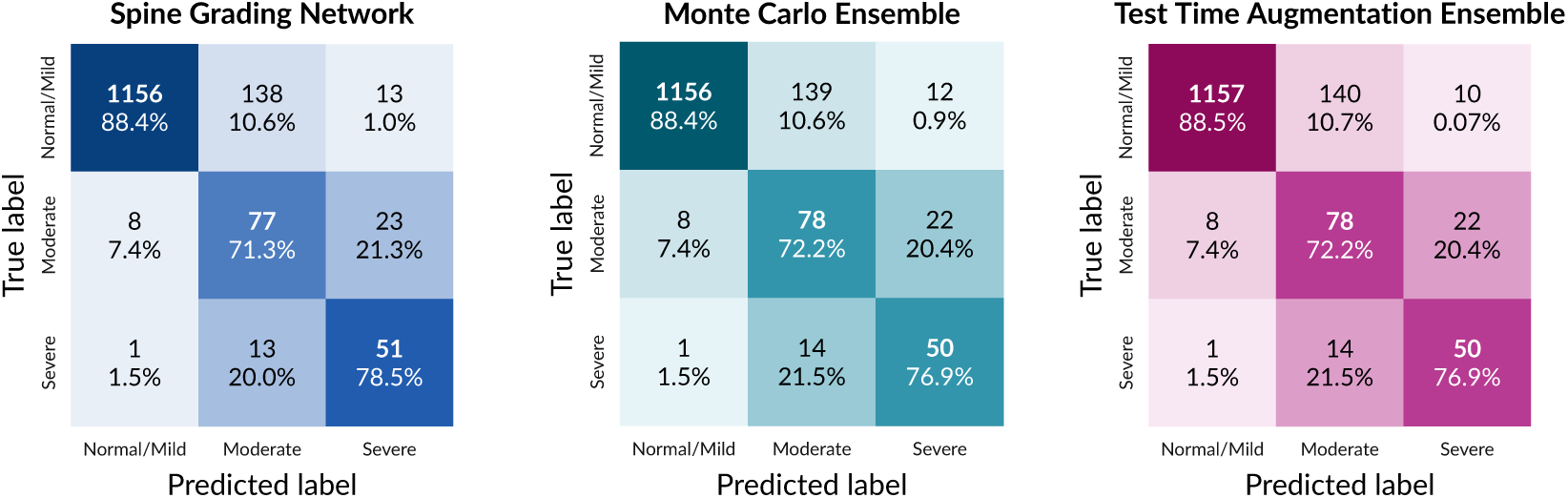
Results for fine-tuned SpineNet Grading Network (SGN) on the test set. Confusion matrix, depicting the number of IVVs and absolute percentages for each class.

## 4 DISCUSSION

This study demonstrates the viability of DL-based automated CCS classification while rigorously addressing critical reliability concerns through uncertainty quantification. Our analysis of five architectures revealed that SGN fine-tuning in the LumbarDISC data set [26] significantly improved diagnostic performance, achieving a balanced accuracy of 79.4%.

The pre-trained SpineNetV2 architecture demonstrated strong performance in identifying normal / mild stenosis cases (98.2% accuracy), but exhibited concerning limitations in more serious conditions, achieving only 20.4% accuracy for moderate and 47.7% for severe stenosis (Table 1). This disparity resulted in a clinically inadequate balanced accuracy of 55.4%, despite the use of class balancing techniques during training [11]. We hypothesize that this reflects a fundamental challenge of generalisation in medical AI, where models trained on a patient population (Genodisc dataset) may struggle when applied to new clinical populations (LumbarDISC dataset) due to differences in imaging protocols, stenosis prevalence, or diagnostic criteria. Incorporating uncertainty estimation could help address this generalisability challenge by providing not only a prediction but also a confidence measure, allowing difficult or out-of-distribution cases to be flagged for further human review.

In addition to SGN, we benchmarked five different model architectures. DenseNet achieved the best performance (69% balanced accuracy), although still significantly below the fine-tuned SGN. Despite the overall good performance of SGN, the confusion matrix revealed persistent challenges in distinguishing moderate from severe cases (21.3% misclassification rate), reflecting known diagnostic difficulties even among human experts [2]. The class imbalance emerged as a critical factor that influences both model performance and uncertainty patterns. After assessing model’s poor calibration using ECE, we applied two well-known methods to estimate uncertainty. Calibration curves and per-class ECE values are provided in Supplementary Fig. 10.

The differences in uncertainty quantification between MC dropout and TTA could be useful in real clinical workflows. MC dropout showed lower uncertainty across all classes, especially for *normal/mild* and *severe* cases. This suggests that its predictions are more stable and could be trusted more when the model is confident. In practice, this means that MC dropout might be helpful to automatically handle mild and severe cases, saving radiologists time by reducing the number of images that need manual review. However, TTA showed higher uncertainty, especially in the *normal/mild* cases. This could actually help spot unusual or borderline normal cases that still deserve a second revision. For example, a case that looks mild but has high uncertainty might reflect an early-stage or a dynamic issue that is not obvious, and this extra flag could prompt additional imaging or follow-up.

For moderate cases - which are often the hardest to classify - both methods resulted in lower confidence, but the uncertainty was still relatively low, especially for MC dropout. This pattern suggests that the model is not confident about the label (low probability), but consistent in that uncertainty. In a clinical setting, these cases could be automatically flagged for further review, helping clinicians focusing on the more challenging diagnoses. In summary, MC dropout could be used to automate decisions on clearer cases, while TTA might be better for identifying subtle or underconfident presentations that need more attention. Using both methods together could support a more balanced and reliable screening process, helping to prioritise the right cases for follow-up or expert review. A minor point worth noting — and a potential limitation — is that we used 25 iterations for MC dropout but only 9 iterations for TTA. This means the uncertainty estimates from MC dropout may be more reliable simply because they are based on more iterations. Furthermore, MC dropout tends to have a smaller effect on the model predictions, since randomly dropping neurons during inference does not drastically alter the output of a well-trained model. In contrast, test-time augmentation can introduce more significant changes, as the input image itself is modified, which can lead to greater variation in predictions. This pattern aligns with the class performance metrics reported in Fig 3, where MC dropout achieved its highest class accuracy for normal/mild cases and relatively lower accuracy for moderate and severe stenosis.

The implementation of uncertainty-aware consensus methods yielded method-dependent improvements. MC dropout with majority voting slightly improved diagnostic consistency in terms of F1 score while maintaining balanced accuracy (77. 4% vs. baseline 79.4%), suggesting clinical viability for multiclass grading. The TTA consensus did not show performance gains, probably due to the inherent variability in the data in the presentation of early stage stenosis that simple augmentation strategies cannot resolve. This suggests that, under TTA, the model is less certain when predicting normal/mild cases compared to more severe categories. In particular, TTA achieved its highest-class accuracy for severe stenosis (84.6%), MC dropout’s consensus predictions improve reliability for multi-class stenosis grading, even with a slight drop in the accuracy. From a deployment perspective, the additional computational cost of uncertainty estimation remains compatible with clinical use. Although stochastic inference increases runtime relative to the base SpineNetV2 model, these absolute processing times remain affordable, and future implementations could further reduce latency through parallelized inference.

Although demonstrating improved reliability in CCS classification, our multicenter retrospective design requires validation across various clinical populations and imaging protocols. In fact, a critical limitation of this study lies in the evaluation of uncertainty estimation exclusively on in-distribution data. The performance and reliability of the model under distribution shift scenarios, common in real-world clinical deployments, remain uncharacterised. Future studies should validate these uncertainty quantification methods on external datasets that exhibit natural variations in imaging protocols and patient demographics. Future work should examine whether predictive uncertainty is consistently higher in misclassified cases, further supporting the clinical utility of uncertainty-based case flagging. Based on previous evidence that demonstrated the superior robustness of deep ensembles to distribution shifts [35], future research should investigate ensemble-based uncertainty frameworks for the prediction of spinal stenosis in heterogeneous clinical environments. Furthermore, to address the higher model confidence observed in majority classes, future studies may explore training strategies to mitigate the effects of class imbalance.

Additionally, to improve clinical trustworthiness, the implementation of post hoc calibration techniques such as temperature scaling [24] may be beneficial. These methods align predicted confidence with actual diagnostic frequencies, essential for assessing difficult cases based on uncertainty thresholds for review by clinicians. Based on our findings, to bridge the gap between AI and clinical application, we could propose a potential clinical workflow in which MC dropout serves as a screening tool to prioritize cases with a high probability of pathology, while cases with high predictive uncertainty are flagged for a more in-depth review by clinicians.

## Data Availability

All data produced in the present work are based on the LumbarDISC
dataset[*] available online .
[*] Richards TJ, Flanders AE, Colak E, Prevedello LM, Ball RL, Kitamura F, et al., The RSNA Lumbar Degenerative Imaging
346 Spine Classification (LumbarDISC) Dataset; 2025. https://arxiv.org/abs/2506.09162.

https://imaging.rsna.org/dataset/6

## Acknowledgements

This project was supported by a grant (# 380, Jutzeler, Manjaly) from the Strategic Focus Area ‘Personalized Health and Related Technologies (PHRT)’ of the ETH domain (Swiss Federal Institute of Technology). The study was also supported by the Swiss National Science Foundation (Ambizione Grant, #PZ00P3_186101, CRJ) and the Schulthess Klinik Research Fund.

For the development of this work, AI-assisted coding systems such as Copilot and Perplexity were used. During the preparation of this manuscript, the author(s) reviewed the text for grammar correctness and improved the content with the help of AI generative tools (Writefull and Perplexity). After using this tool/service, the author(s) reviewed and edited the content as needed and take(s) full responsibility for the content of the publication.

## Conflict of interest

CRJ serves as a scientific consultant for Abbvie and Mitsubishi Takeda; however, this role did not influence the design, conduct, or reporting of this study. All other authors declare no conflict of interest.

## Author Contribution

A.B.: Conceptualization, Data curation, Formal analysis, Investigation, Methodology, Software, Visualization, Writing – review & editing. M.M.: Conceptualization, Formal analysis, Investigation, Validation, Methodology, Project administration, Software, Visualization, Writing – original draft, Writing – review & editing. F.G.: Supervision, Writing – review & editing. Z.M.M.: Conceptualization, Funding acquisition, Resources, Writing – review & editing. A.C.: Conceptualization, Methodology, Supervision, Visualization, Project administration, Validation, Writing – original draft, Writing – review & editing. C.R.J.: Funding acquisition, Writing – review & editing.

## Supporting Information

This appendix provides a comprehensive overview of the CCS grading using the deep learning models hyperparameter optimization process, benchmarking strategy, and detailed classification performance analysis.

### CCS classification model and hyperparameter optimization and benchmarking

Building reliable deep learning models for CCS classification involves systematic evaluation across various architectures and optimizing training parameters. With significant class imbalance in stenosis datasets (88% normal/mild, 7% moderate, 5% severe), our strategy focuses on balanced accuracy to ensure consistent performance across all severity levels.

To optimize the performance of the models, we conducted a comprehensive hyperparameter random search, testing different batch sizes (16, 32) and learning rates (10^−3^ to 10^−6^) with variable dropout rates (0-0.9 in 0.1 steps). Models were trained for 30-130 epochs with early stopping after 20 epochs if there was no improvement in validation loss. Warm-up and cool-down mechanisms were used to stabilize training and enhance convergence. The warm-up was linear over 1000 batches with a factor of 0.01. Cooldown reduced the learning rate by 0.5 on a plateau if validation loss did not improve after 5 epochs, using a minimum learning rate of 10^−7^. To improve training and model generalisation, two data augmentations were applied with a probability of 0.5, allowing multiple transformations per sample: random zooming from 90 to 110%, maintaining IVD volume size, and random Gaussian noise with a mean of 0 and a standard deviation of 0.01. We selected the optimal set of hyperparameters based on the balanced accuracy performance in the validation data set.

The hyperparameter optimization process was carried out through a systematic random search to maximize the balanced accuracy at all stenosis severity levels. Although DenseNet employed a weighted random sampler strategy, all other models utilized weighted cross-entropy to address class imbalance. Table 2 presents the final hyperparameter configuration for each model architecture evaluated in this study.

**TABLE 2.** Hyperparameters of the model architectures with the best performance according to the balanced accuracy.

| Parameters | fine-tuned SGN | DenseNet | ResNet | EfficientNet |
| --- | --- | --- | --- | --- |
| Batch size | 16 | 16 | 16 | 16 |
| Learning rate | $10^{-6}$ | $10^{-3}$ | $10^{-4}$ | $10^{-4}$ |
| Weighted random sampler | False | True | False | False |
| Weighted Cross Entropy | True | False | True | True |
| Dropout rate | 0.8 | 0.2 | 0.2 | 0.3 |

Five deep learning architectures were benchmarked for their ability to classify CCS severity. Figure 6 shows confusion matrices that highlight key insights into each model’s classification behavior. The confusion matrices reveal consistent interclass confusion between moderate and severe stenosis in all architectures, with misclassification rates ranging from 17.6% to 23.1%.

**FIGURE 6.**
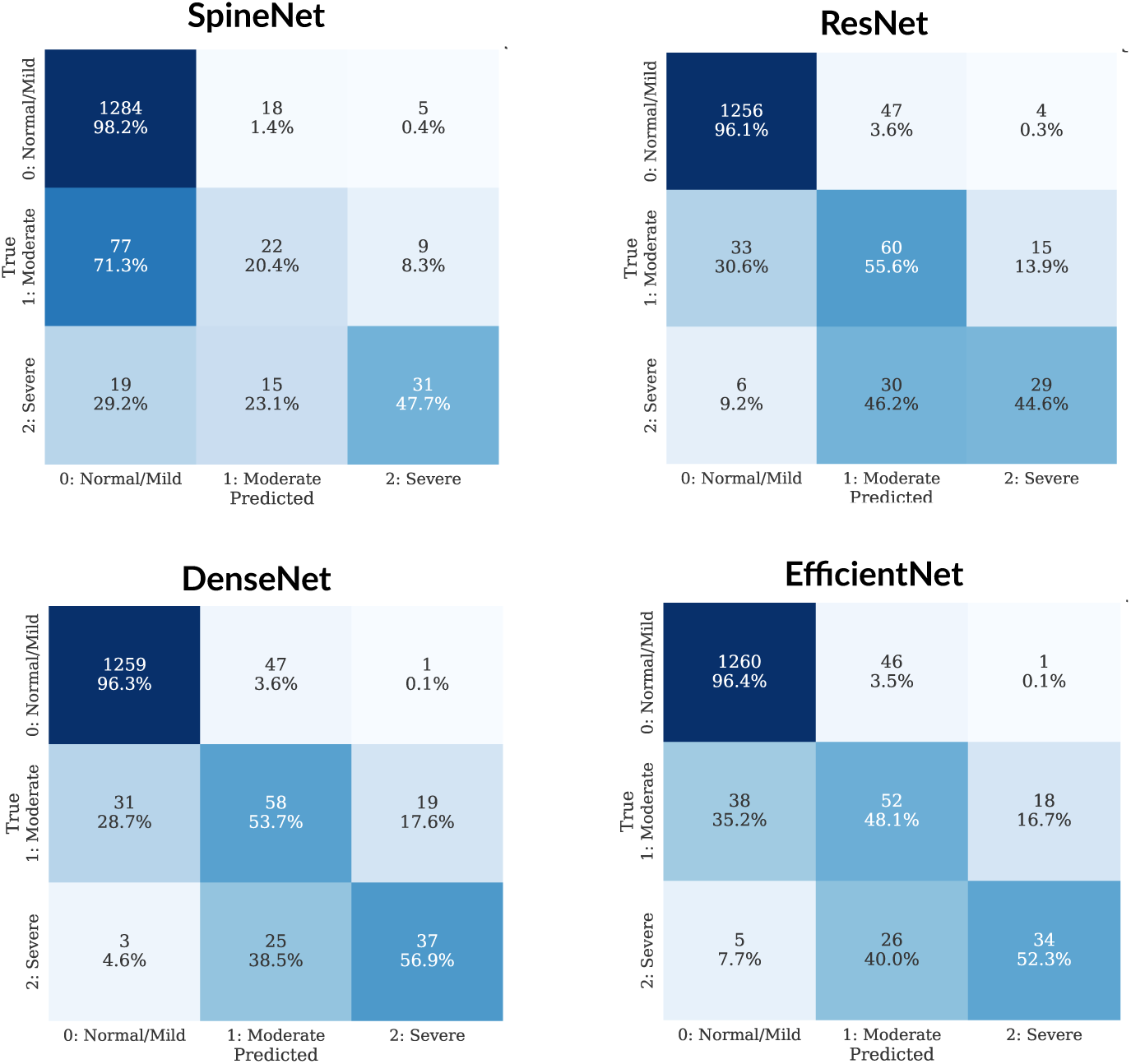
Confusion matrices for different deep learning architectures on lumbar central canal stenosis classification. Results show model classified samples across *Normal/Mild, Moderate*, and *Severe* stenosis classes.

DenseNet, ResNet, and EfficientNet demonstrated moderate performance, while original SpineNetV2 showed limited generalisability to the target dataset. DenseNet achieved the second highest performance with a balanced accuracy of 69. 0% and a competitive macro F1 score of 68.7%. ResNet and EfficientNet exhibited moderate performance levels, with both models achieving similar balanced accuracy (65.4% and 65.6%, respectively) and macro F1 scores (65.6% and 66.3%, respectively). In contrast, the original SpineNet architecture demonstrated a significant performance limitation, achieving high accuracy (98.2%) for the majority normal/mild stenosis class but showing poor performance for minority classes, with only 20.4% accuracy for moderate stenosis and 47.7% for severe stenosis, resulting in an overall balanced accuracy of just 55.4%.

### Test Time Augmentations

The implementation of TTA used nine image perturbations the model’s sensitivity to controlled input variation during inference, rather than to simulate anatomically realistic MRI acquisitions. Figure 7 illustrates the nine image perturbations used during TTA inference.

**FIGURE 7.**
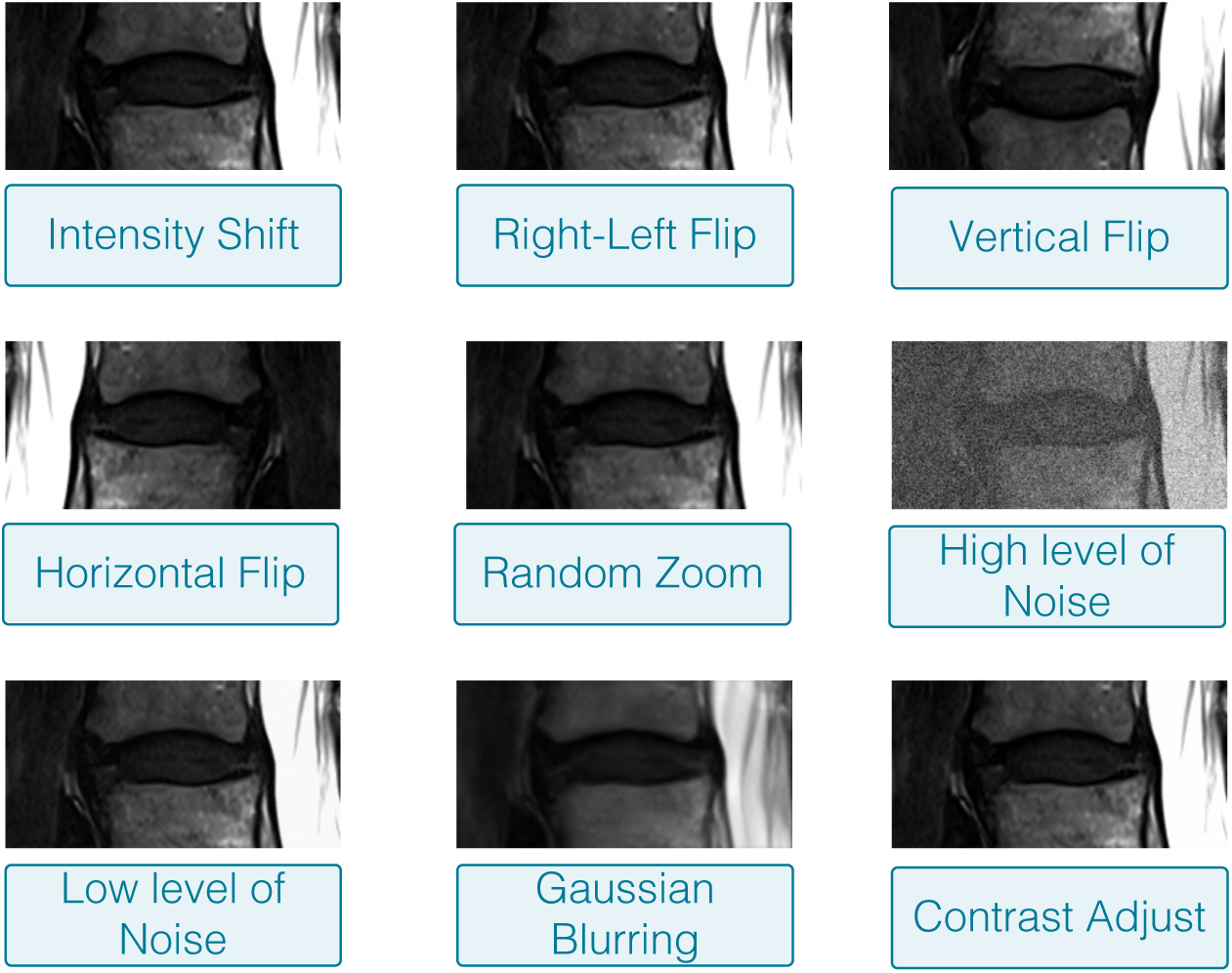
Visualization of image augmentation transformations applied for Test Time Augmentation (TTA)

- Intensity shift: random intensity offset in the range [-0.05, 0.05] to perturb global signal levels
- Contrast Adjustment: gamma correction with values between 0.95 and 1.05 to perturb image contrast
- Gaussian Noise Addition (low-level): noise perturbation with standard deviation of 0.01 to introduce a mild noise perturbation
- Gaussian Noise Addition (high-level): noise perturbation with standard deviation of 0.2 to introduce a strong noise perturbation
- Right-left flip: image reflection used as a geometric perturbation
- Vertical Flip: image reflection used as a geometric perturbation
- Horizontal Flip: image reflection used as a geometric perturbation
- Random Zoom: scale variation between 90% and 110% while preserving the final intervertebral disc volume dimensions
- Gaussian Blurring: smoothing perturbation to reduce local high-frequency detail

### Uncertainty Estimation

A comprehensive comparative visualization of uncertainty estimates between Monte Carlo (MC) dropout and Test Time Augmentation (TTA) methods across all stenosis severity classes is presented in Figure 8. The stratified approach assesses each uncertainty quantification method when the true diagnosis is known.

**FIGURE 8.**
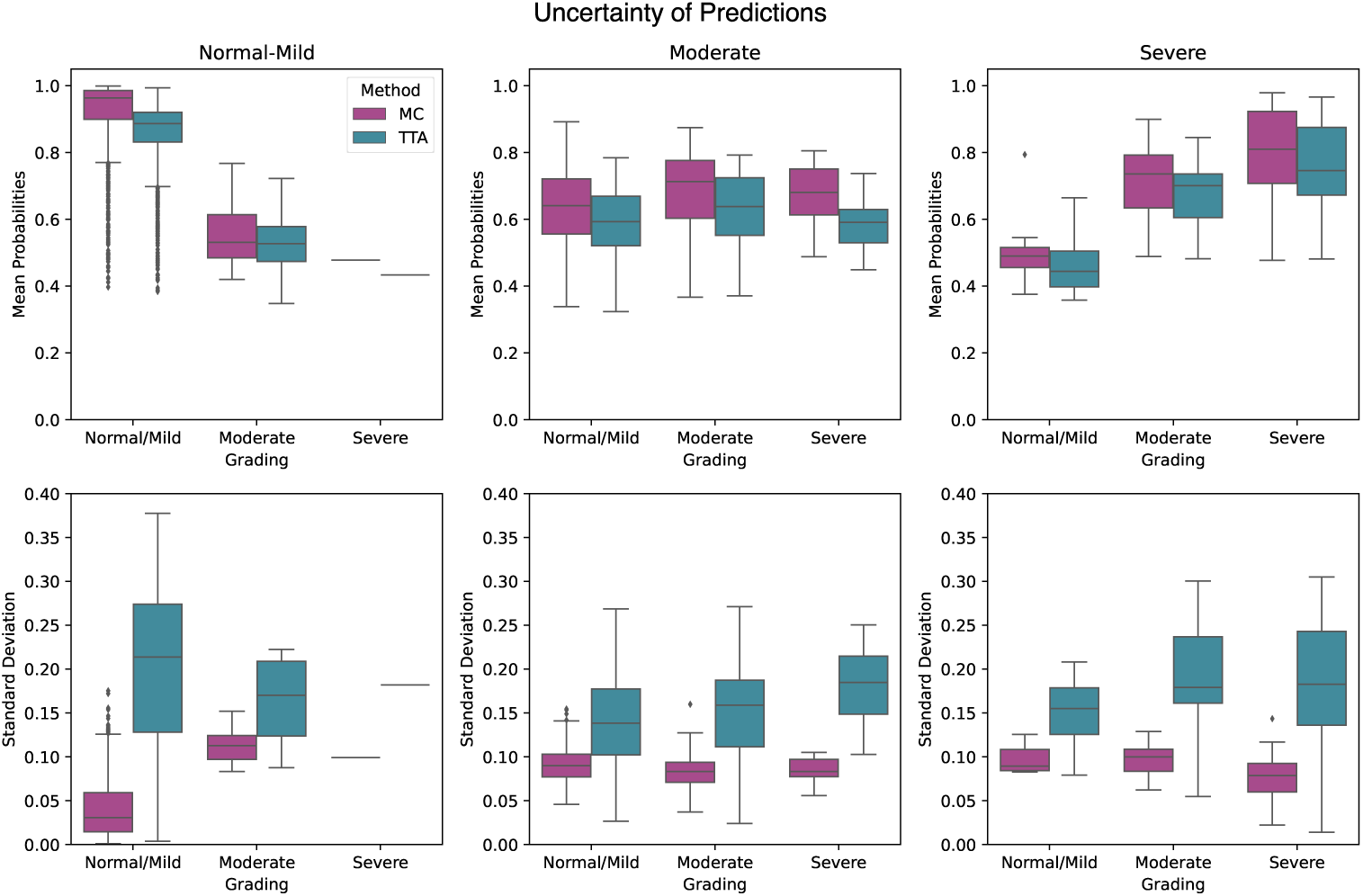
Box plots show mean predicted probabilities (top row) and standard deviations for predicted severity class (Normal/Mild, Moderate, Severe) using Monte Carlo dropout (MC) and Test Time Augmentation (TTA).

Figure 8 shows uncertainty patterns for predicted classes, with results organized by true stenosis severity, offering insights into model confidence and reliability across categories. MC dropout maintains high confidence across all classes, with median probabilities of 0.963 for Normal/Mild, 0.673 for Moderate, and 0.762 for Severe stenosis, indicating strong epistemic uncertainty quantification. Uncertainty, measured by standard deviations, increases with stenosis severity: Normal/Mild (median 0.031, IQR 0.014-0.060), Moderate (median 0.087, IQR 0.074-0.099), Severe (median 0.088, IQR 0.070-0.104). This trend matches clinical expectations, as complex pathologies introduce more ambiguity. TTA shows lower confidence with median probabilities of 0.887 for Normal/Mild, 0.605 for Moderate, and 0.701 for Severe stenosis, reflecting sensitivity to data variation and capturing aleatoric uncertainty. Uncertainty is highest in Normal/Mild cases (median 0.213, IQR 0.128-0.273), followed by Severe (median 0.179, IQR 0.134-0.233), and lowest in Moderate (median 0.148, IQR 0.108-0.185). This suggests TTA is sensitive to early pathology variations and severe distortions. MC dropout has narrow uncertainty distributions, most spread in Moderate and Severe cases (IQR 0.025 and 0.034), while TTA’s broader distributions, especially in Normal/Mild cases (IQR 0.145), show more sensitivity to input changes.

### Predictive entropy and classification errors

To complement the standard deviation-based analysis, we evaluated predictive entropy across repeated model predictions. For each intervertebral volume, MC Dropout and TTA generated multiple class-probability estimates, which were averaged to obtain a final probability profile for each case. Predictive entropy was then calculated from this average distribution. For each input sample, the model produced *T* softmax probability vectors, where *T* = 25 for MC Dropout and *T* = 9 for TTA. Let *p* ^(^*^t^* ^)^ ^∈R^*^K^* denote the softmax output at stochastic pass *t*, with *K* = 3 classes. The mean predictive distribution was then computed as

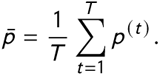

Predictive entropy was calculated from this averaged distribution as

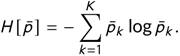

This quantity summarizes the total uncertainty of the predictive distribution and complements the standard deviation of the predicted class probability used in the main analysis. Predictive entropy reflects how clearly the model favours one stenosis category over the others. Lower entropy indicates a more concentrated prediction, whereas higher entropy reflects greater ambiguity in the final class assignment. As shown in Supplementary Figure 9, mis-classified Normal/Mild and Severe cases had significantly higher entropy than correctly classified cases for both MC Dropout and TTA. No significant difference was observed for Moderate stenosis, consistent with the greater ambiguity of this intermediate class and the overlap between adjacent grades. Overall, these findings indicate that higher predictive entropy is associated with a higher likelihood of classification error in the more clearly separable classes.

### Calibration analysis

Calibration was further assessed using Expected Calibration Error (ECE). Figure 10 shows that MC dropout was better calibrated overall than TTA, although both methods displayed residual class-specific miscalibration. These findings further support the use of explicit uncertainty estimates in addition to raw predicted probabilities.

**FIGURE 9.**
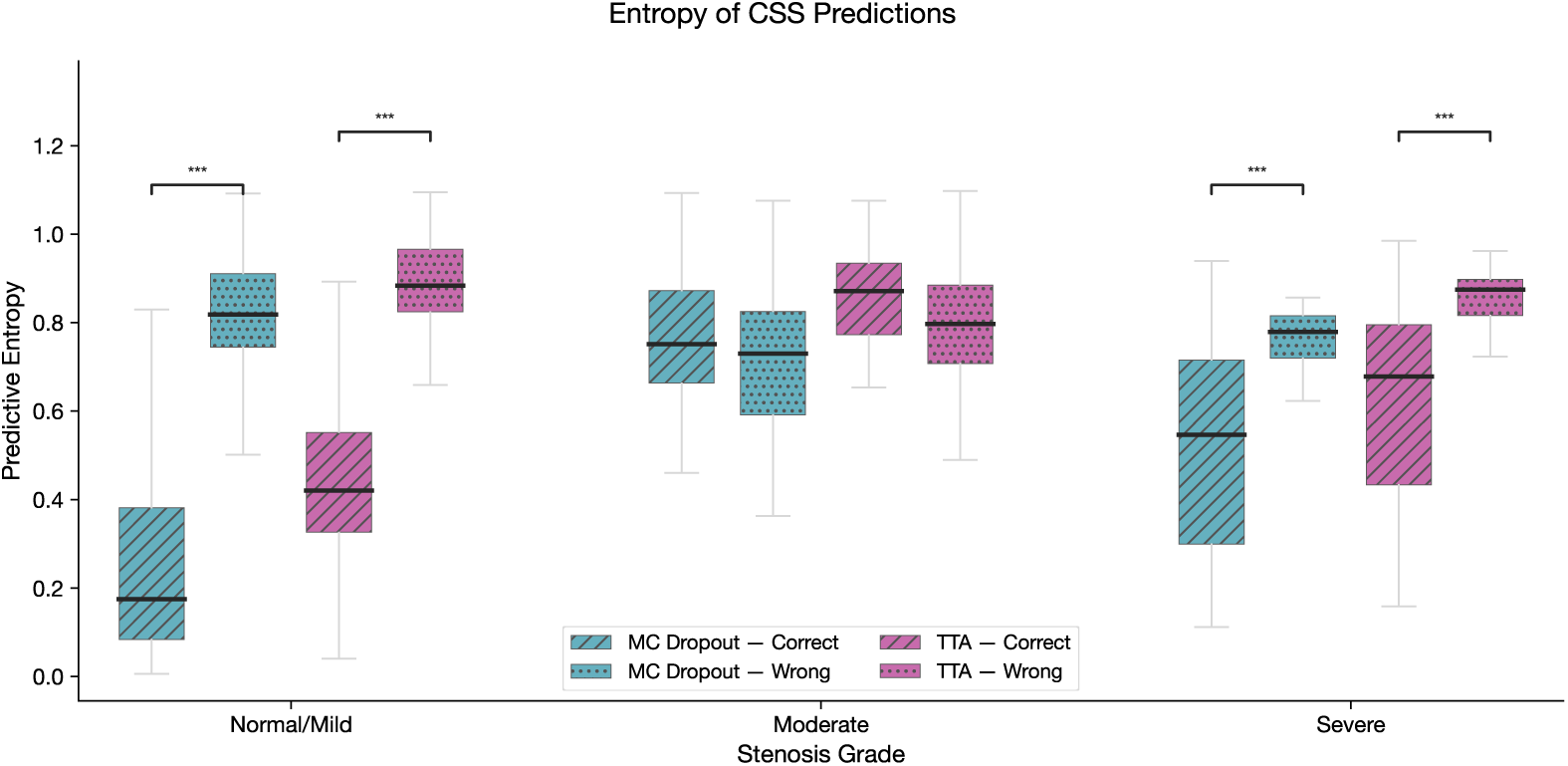
Predictive entropy of CCS grading predictions, stratified by classification outcome, for MC Dropout (*T* =25 passes) and Test-Time Augmentation (TTA, *T* =9 passes). For each spine level, repeated stochastic predictions generated class-probability vectors which were averaged to obtain the final predictive distribution. Higher entropy indicates greater uncertainty in the final stenosis grade, whereas lower entropy indicates a more focused and confident class assignment. Boxes show the distribution of for correctly classified and misclassified spine levels within each stenosis grade. Brackets indicate statistically significant differences between correct and incorrect predictions (one-sided Mann–Whitney U test, ^∗∗∗^*p* < 0.001). Misclassified Normal/Mild and Severe cases showed significantly higher entropy under both methods, whereas no significant difference was observed for Moderate stenosis, consistent with the greater ambiguity of this intermediate class.

**FIGURE 10.**
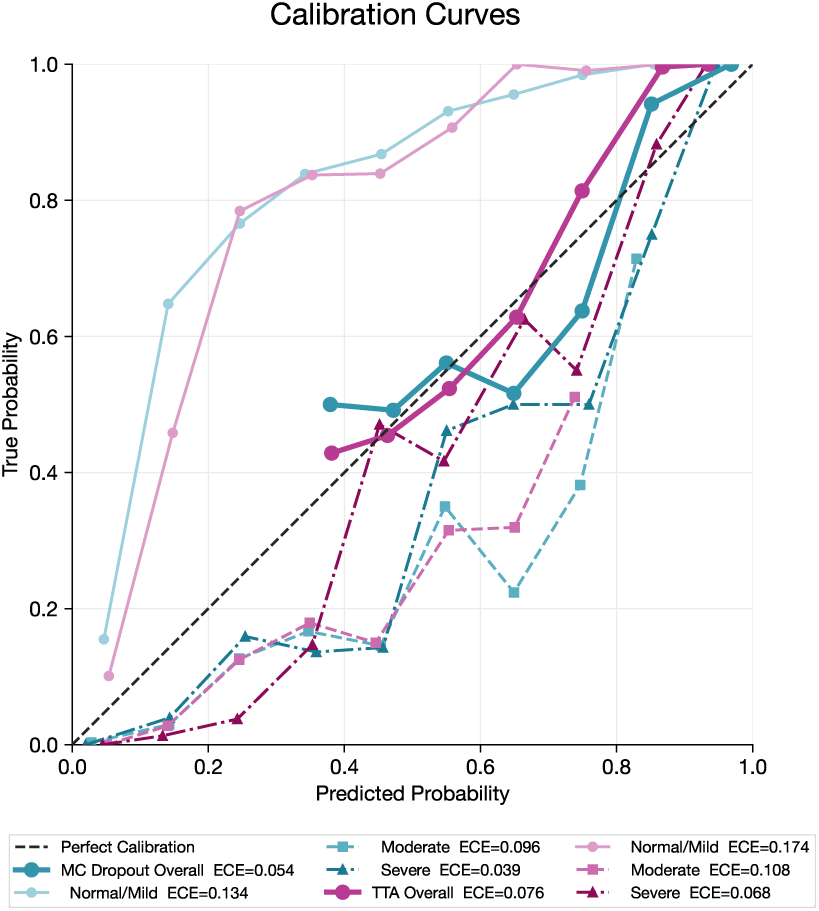
Reliability curves and Expected Calibration Error (ECE) for Monte Carlo dropout (MC) and Test Time Augmentation (TTA). Lower ECE indicates better agreement between predicted confidence and empirical accuracy.

## Notes

### Competing Interest Statement

The authors have declared no competing interest.

### Summary of Updates

THe Figure were revised and the entropy was also computed

